# Automatic framework for evaluating osteoarthritic cartilage severity: high-resolution cartilage thickness mapping and scoring

**DOI:** 10.1101/2025.11.03.25339377

**Authors:** Paul Margain, Patrick Omoumi, Julien Favre

## Abstract

**Objectives:** To develop and validate an automatic, scalable framework for assessing the femoro-tibial osteoarthritic cartilage severity using high-resolution cartilage thickness maps (CTh-Maps) and a cartilage thickness scoring system (CTh-Score).

**Methods:** The Osteoarthritis Initiative (OAI) cohort of 4796 subjects was analyzed. A 3D-UNet was trained to segment femoro-tibial bones and cartilages using MRI from baseline, 1-, 2-, 3-, 4-, 6- and 8-year follow-ups. CTh-Maps were created for each knee. A ResNet model trained on CTh-Maps assigned a CTh-Score ranging from 0 (healthy cartilage) to 100 (end-stage OA). The reproducibility of the CTh-Score was evaluated in a test/retest setup. Its validity was assessed by examining the correlation with expert evaluations of cartilage loss (MOAKS grading) and association to OA severity (KL grade) in both OAI and external dataset. The CTh-Score sensitivity to OA structural progression was examined.

**Results:** The framework generated CTh-Maps for the entire OAI, forming the “OAI CTh-Maps” dataset. Both CTh-Maps and CTh-Score showed excellent reproducibility (ICC>0.98). The CTh-Score demonstrated strong correlations (r=0.81) with expert assessments of cartilage loss and strong associations to OA severity, including in the external dataset. The CTh-Score either increased or remained stable for almost all subjects at 8-year follow-up. The CTh-Score showed great sensitivity to change, significantly increasing between each timepoint, up to 6 years prior to KL progression.

**Conclusions:** CTh-Maps and CTh-Score represent a novel approach to analyze cartilage at imaging. Their scalability, reproducibility and sensitivity to osteoarthritic cartilage severity provide significant opportunities for earlier OA detection, better disease monitoring, and therapeutic window identification.

## Introduction

Although osteoarthritis (OA) is now considered a whole-organ disease, cartilage degeneration represents the primary hallmark of the condition. Cartilage degeneration progresses throughout all stages of the disease, encompassing a wide range of changes. Initially, this may start with potential swelling, followed by varying degrees of cartilage loss from surface fibrillation to full-thickness substance loss[1]. The extent of these changes can also differ across the cartilage surfaces. Magnetic Resonance Imaging (MRI) has been the preferred method for non-invasive assessment of structural changes in cartilage [2], [3] providing quantitative assessment of cartilage thickness (CTh). The analysis of CTh has revealed that OA is not a simple, one-directional process towards homogeneous cartilage substance loss[4]. Instead cartilage degeneration follows diverse CTh patterns, including both areas of thinning and occasional thickening across joint surfaces, with varying spatial distribution between subjects[5], [6].

To facilitate comparisons, CTh measurements commonly rely on region-of-interest (ROI)-based approaches, which provide useful insights into CTh changes but exhibit weak correlations with disease status[7]. This limitation arises from two main factors. First, the average CTh in ROIs for individuals with OA largely overlaps with that of healthy subjects. Second, across OA stages, ROI averaging fails to capture the full complexity of lesion patterns because of their limited spatial resolution, resulting in a loss of critical spatial information.

To address this limitation, CTh-Maps have been introduced as high-resolution “thickness images” that capture femoro-tibial CTh measurements across the entire articular surface [5], [8]. These images are anatomically standardized, ensuring pixel-to-pixel correspondence for inter-subject comparisons. Generating CTh-Maps requires precise segmentation across all slices. Initially, this was achieved through manual segmentation, which is considered the gold standard for precision but is also extremely time-consuming. To adequately capture the full diversity of cartilage degeneration patterns, it is essential to analyze large cohorts of MRI data due to their inherent variability. This necessity underscores the importance of developing an automatic and reliable pipeline for processing CTh-Maps, facilitating efficient analyses at large-scale.

Deep learning methods are particularly relevant for both processing and analyzing CTh-Maps. First, recent advancements have enabled precise automatic segmentation, addressing the longstanding challenge of segmenting small tissues with complex anatomy such as cartilage[9]. Second, convolutional neural networks (CNN) [10] offer a powerful approach for analyzing high-resolution CTh-Maps. By applying convolutional filters that slide across an image, CNN effectively capture local features and learn spatial patterns. Their robustness to diverse lesion patterns has been demonstrated in numerous medical imaging applications, making them well-suited for identifying and characterizing CTh degeneration patterns [10].

A key advantage of CNN is their ability to learn meaningful representations even from noisy or imperfect training data[11]. In the absence of a dedicated ground truth for osteoarthritic cartilage severity, CNN can be trained using a weakly supervised approach with the widely used radiographic global OA severity, the Kellgren-Lawrence grade. By leveraging large and diverse datasets, CNN has the potential to learn to score the CTh patterns with respect to disease severity, hereby providing a tool to quantitatively assess osteoarthritic cartilage severity for a given knee.

Therefore, this study aimed to:

1. Develop an automatic framework for generating CTh-Maps and assess its reliability.
2. Establish a scoring system (CTh-Score) to quantify osteoarthritic cartilage severity based on CTh-Maps and evaluate its reliability and validity.

## Materials and Methods

### Patient population and data extraction

The OAI [12], [13] is a large-scale, multicenter, longitudinal, and prospective observational study designed to investigate knee osteoarthritis. The study includes 4,796 participants who either have or are at risk of developing knee OA. Imaging data were collected at baseline (00m) and follow-up visits conducted at 12, 24, 36, 48, 72, and 96 months. The dataset is publicly available online. The imaging dataset comprises high-resolution sagittal 3D dual-echo at steady-state water-excitation (DESS) MRI and standardized knee radiographs. All imaging procedures adhered to a standardized acquisition protocol [12], [13], [14].

MRI and radiographic readings were extracted from the OAI repository. The Kellgren-Lawrence grade (KL grade) [15], which serves as the reference standard for assessing global structural OA severity in radiography, ranging from 0 (no radiographic OA) to 4 (severe OA), was extracted for all knees at all timepoints. The MRI Osteoarthritis Knee Score (MOAKS) [15] is a semi-quantitative grading system used for whole-joint assessment, including partial/full cartilage loss. Within the tibiofemoral joint, cartilage loss is graded on a scale from 0 to 3 based on the size of the lesion relative to the affected region. Since MOAKS assesses cartilage loss within specific regions of interest, the gradings for the size of cartilage loss (indicated by the first digit in MOAKS cartilage assessment) were summed across all ROIs to create a comprehensive measure for expert evaluations of total size of cartilage loss in correlation analyses. MOAKS data were extracted for a sub cohort of the OAI at baseline.

### Cartilage thickness maps dataset

#### Segmentation (Figure 1 – Step A)

For each available DESS sequence, automatic segmentation of the femoro-tibial bone and cartilage was performed using a 3D-UNet model trained with 5-fold cross-validation using nnUNet framework [16], [17]. The network was trained on 507 manual segmentations, which have been validated in prior studies to develop and evaluate automatic segmentation methods for the OAI dataset[18], [19].

**Figure 1.**
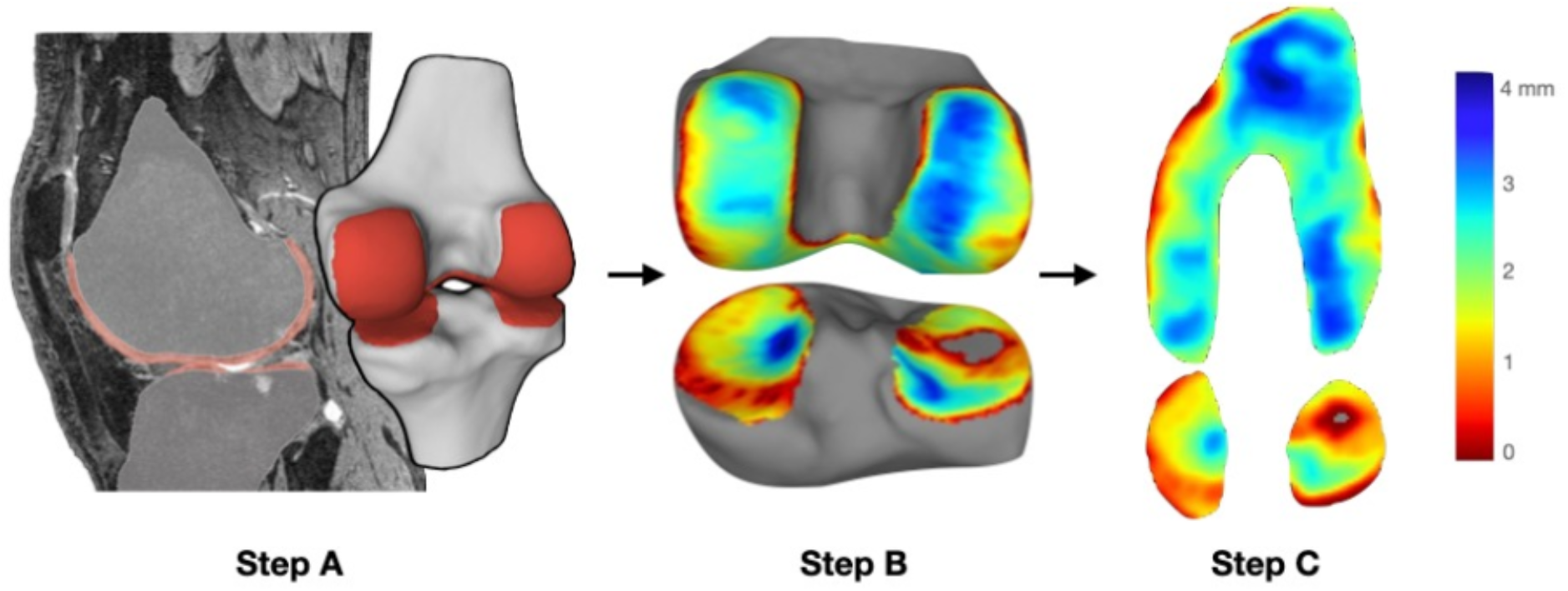
Automatic pipeline for cartilage thickness map generation. Steps of processing: segmentation (A), anatomical standardization (B), and 2D transformation (C). (A) the MRI is automatically segmented resulting in bone (grey) and cartilage (red) masks of femur and tibia. (B) The masks are converted into meshes and an atlas bone is registered. The cartilage thickness is calculated in the subchondral vertices as the distance to the cartilage surface. (C) 3D subchondral surface is transformed into a 2D high resolution images of cartilage thickness.

#### Standardizations (Figure 1 – Step B & C)

After segmentation, each bone and cartilage masks were converted to 3D surface triangular meshes[20]. An atlas mesh, containing bones and subchondral delimitations that were created by averaging healthy bones, was reused from a previous study[21]. This atlas was registered to each femur and tibia bone meshes separately[22], [23]. Cartilage thickness was calculated as the surface-to-surface distance between the vertices of the subchondral bone of the registered atlas mesh and the cartilage surface. Full cartilage loss was detected when bone surface normal did not intersect the cartilage mesh; in such cases, a value of zero was assigned for cartilage thickness[19]. The 3D anatomically standardized cartilage thickness maps were expressed as 2D maps using cylinder mapping for the femur and projection for the tibia[5].

### CTh-Score

To quantify cartilage thickness patterns, a convolutional neural network (ResNet18) was trained on OAI baseline CTh-Maps. The CTh-Maps were converted to three-channel inputs by duplicating the single-channel maps. A separate test set consisting of 600 subjects was reserved to evaluate the model’s generalizability. In a supervised regression task, a CNN is trained to recognize patterns in input images that correspond to a target output. While no definitive reference exists for scoring cartilage degenerations, cartilage damage generally worsens with increasing structural disease severity. Therefore, the network’s target was set to the global structural severity, represented by the KL grade[24], [25]. By leveraging the large OAI dataset, the model was trained to identify cartilage-specific lesion patterns associated with disease severity. The network’s output was linearly rescaled to a 0–100 scale, yielding the Cartilage Thickness Score (CTh-Score). This score reflects disease severity, where 0 corresponds to healthy cartilage and 100 indicates advanced cartilage lesions. Figure 2 illustrates the CTh-Maps and the corresponding CTh-Scores across the seven timepoints of the OAI dataset for five representative knee subjects.

**Figure 2.**
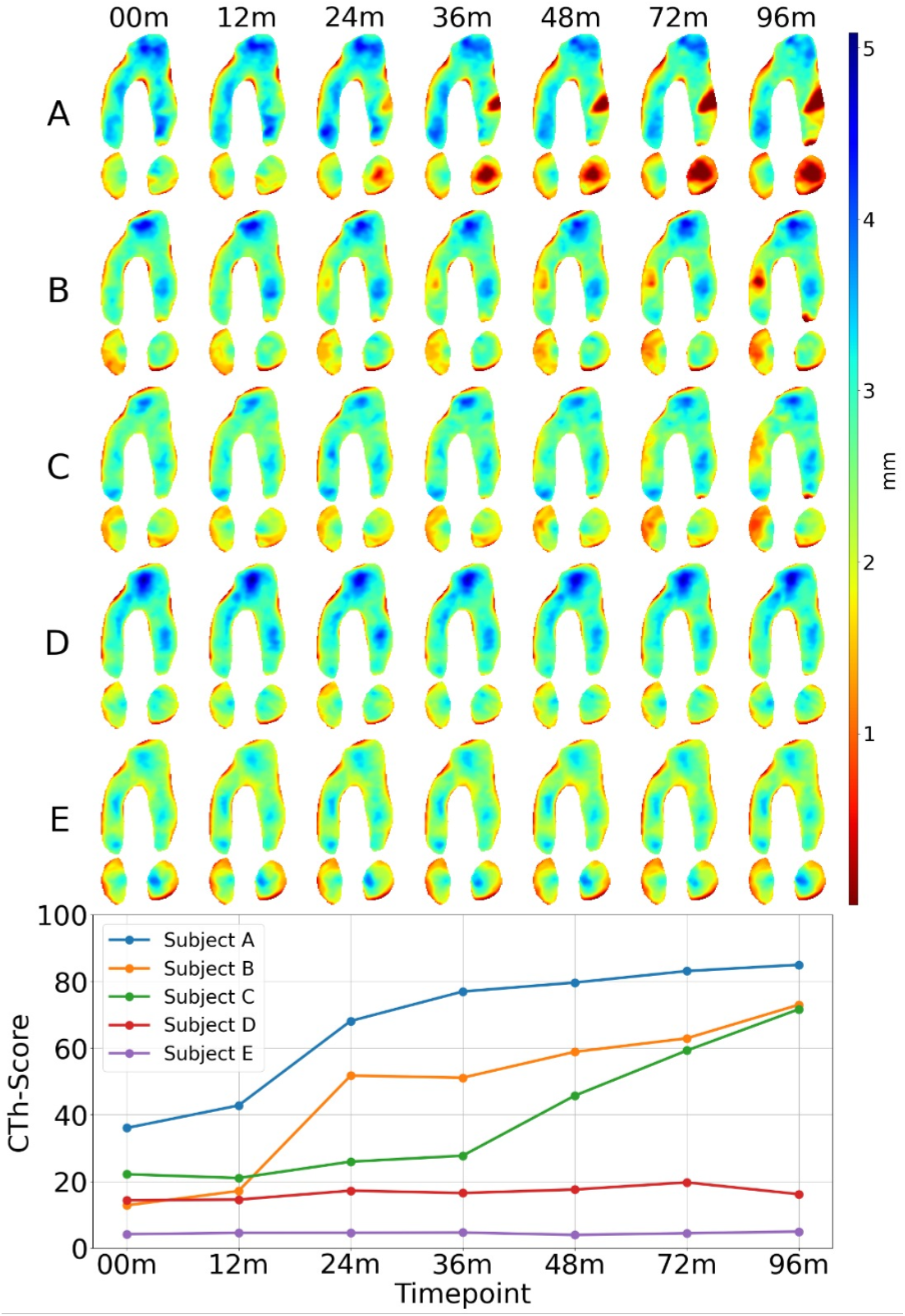
Five representative subject knees longitudinal CTh-Maps and corresponding CTh-Scores. Subject A presents cartilage loss in the lateral femur and tibia. Subject B and C present cartilage loss in the medial femur and tibia. Subject D and E apparently do not present cartilage loss. One can qualitatively appreciate the subject specific patterns of CTh changes on CTh-Maps and the ability of the CTh-Score to evaluate the lesions independently of their location.

### Repeatability of CTh-Maps and CTh-Score

To test the repeatability of CTh-Maps and CTh Scores, the OAI pilot study images were used. This dataset includes 19 subjects (12 women; 10 with OA) who underwent test-retest examinations of their left or right knee. OA participants were selected using the OAI study design and documentation. The mean age was 51 years (range 40–71), and the mean BMI was 30.4 kg/m^2^ (range 19.1–44.0).

For the CTh-Maps, the mean and standard deviation of errors (differences between first and second imaging/processing) defined the accuracy and precision of reproducibility, respectively[8]. Agreement was evaluated using a two-way random-effects intraclass correlation coefficient (ICC) between map pixels from first and second imaging/processing[8].

For the CTh-Score, repeatability was assessed by calculating the ICC and the smallest detectable difference (SDD), defined as the threshold below which differences cannot be reliably distinguished from random measurement error. The SDD was calculated using the 95% confidence interval of the error standard deviation (limits of agreement method of Bland and Altman)[26].

### Generalizability of CTh-Score

To assess the generalizability of the CTh-Score (i.e. its robustness to out-of-training data), the network trained on OAI CTh-Maps to produce the CTh-Score was tested on the internal test set and on an independent external dataset from a previously published study [27]. This dataset consisted of knee CT arthrograms, covering KL grades 0 to 4 (n = 110, 41, 45, 62, 32), obtained in a different patient population with previously published description [27]. Manual ground-truth segmentations for bone and cartilage were available for the CT-arthrography dataset, and we retrained the segmentation model and verified high segmentation accuracy (Dice > 0.9). Segmentations from this dataset were processed using the same pipeline to generate CTh-Maps. Without additional retraining, the trained network assigned CTh-Scores to these new CTh-Maps. To evaluate the CTh-Score’s ability to reflect cartilage status in relation to OA severity, statistical comparisons were conducted using t-tests between CTh-Scores corresponding to consecutive KL grades.

### Validity of CTh-Score

#### CTh-Score trajectories in the OAI cohort over 8 years

In OA disease course, the cartilage may exhibit increasing or stable severity. In figure 4, for each participant’s knee, the evolution of CTh-Score from baseline to the last imaging follow-up (8 years later) was examined to verify these trends. The distribution of CTh-Score, increasing as a function of the baseline CTh-Score, was also reported.

**Figure 3.**
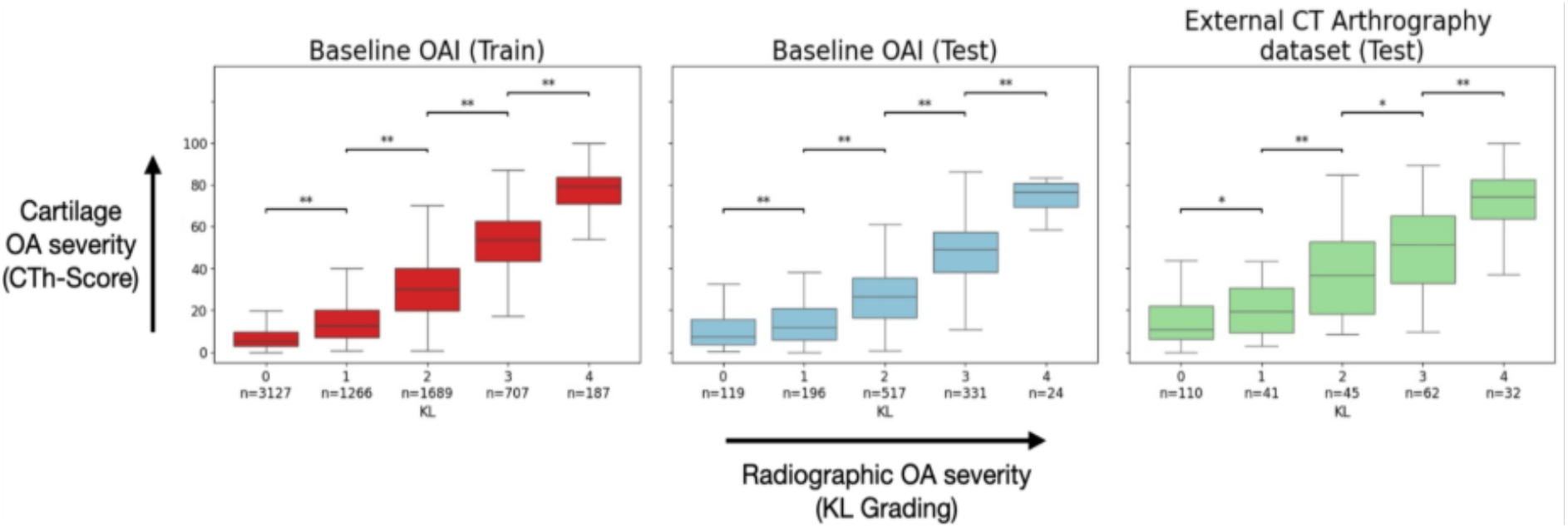
CTh-Score demonstrates sensitivity to radiographic OA severity and generalizability across datasets. The distribution of the CTh-Score is shown stratified by Kellgren-Lawrence (KL) grade for: **(left)** OAI baseline training set, **(middle)** OAI baseline test set, and **(right)** External CT arthrography validation set. The CTh-Score significantly increases between successive KL grades (** p<0.001, * p<0.05) in all cohorts. This consistent ability to distinguish between KL grades, maintained in the test and external validation sets, confirms the score’s sensitivity to radiographic severity and its robustness across unseen data and differing imaging setups.

**Figure 4.**
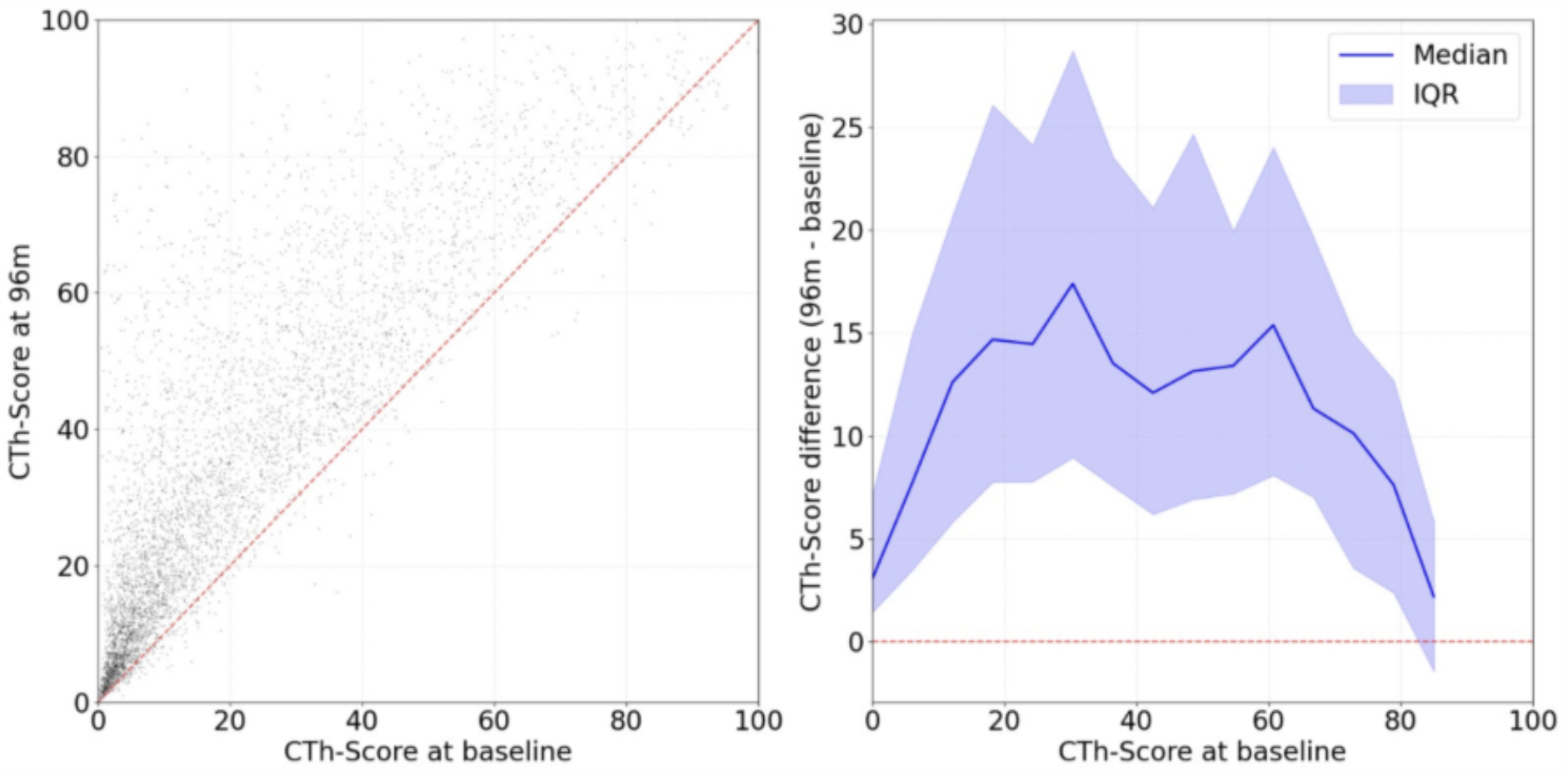
Eight-year progression of CTh-Scores (left). Distribution of CTh-Score difference (CTh-Score_96m_ – CTh-Score_baseline_)(right) in the entire OAI cohort. The vast majority of subject knees experienced an increase in their CTh-Score. While a high variability is observed, the magnitude of this increase generally follows a pattern: it initially rises for CTh-Score_baseline_ between 0 and 20 (early lesions), plateaus between 20 and 60 (active lesions), and then gradually declines beyond 60 (reflecting severe lesions that have limited potential for further worsening).

#### Relationship with cartilage loss and global disease severity

To evaluate the relationship between the CTh-Score and cartilage loss, the expert assessments of total size of cartilage loss from MOAKS gradings in the OAI MOAKS subcohort were correlated with the CTh-Score. Additionally, correlations with KL grade were also calculated to assess the added capability of the CTh-Score to reflect global disease severity.

#### Sensitivity to disease progression

Figure 5 examines how the continuous nature of the CTh-Score, unlike the KL’s five discrete grades, enhances the ability to detect early changes in OA cartilage severity prior to any KL grade progression. Participants who experienced a KL grade increase were categorized into four transition groups (corresponding to the timepoint where the KL grade progresses from one grade to the next). The CTh-Score was then assessed at multiple time points (6 years, 4 years, and 2 years prior to KL grade progression, as well as at the time of transition) to determine whether it could detect disease progression before it became radiographically apparent through the KL grade.

**Figure 5.**
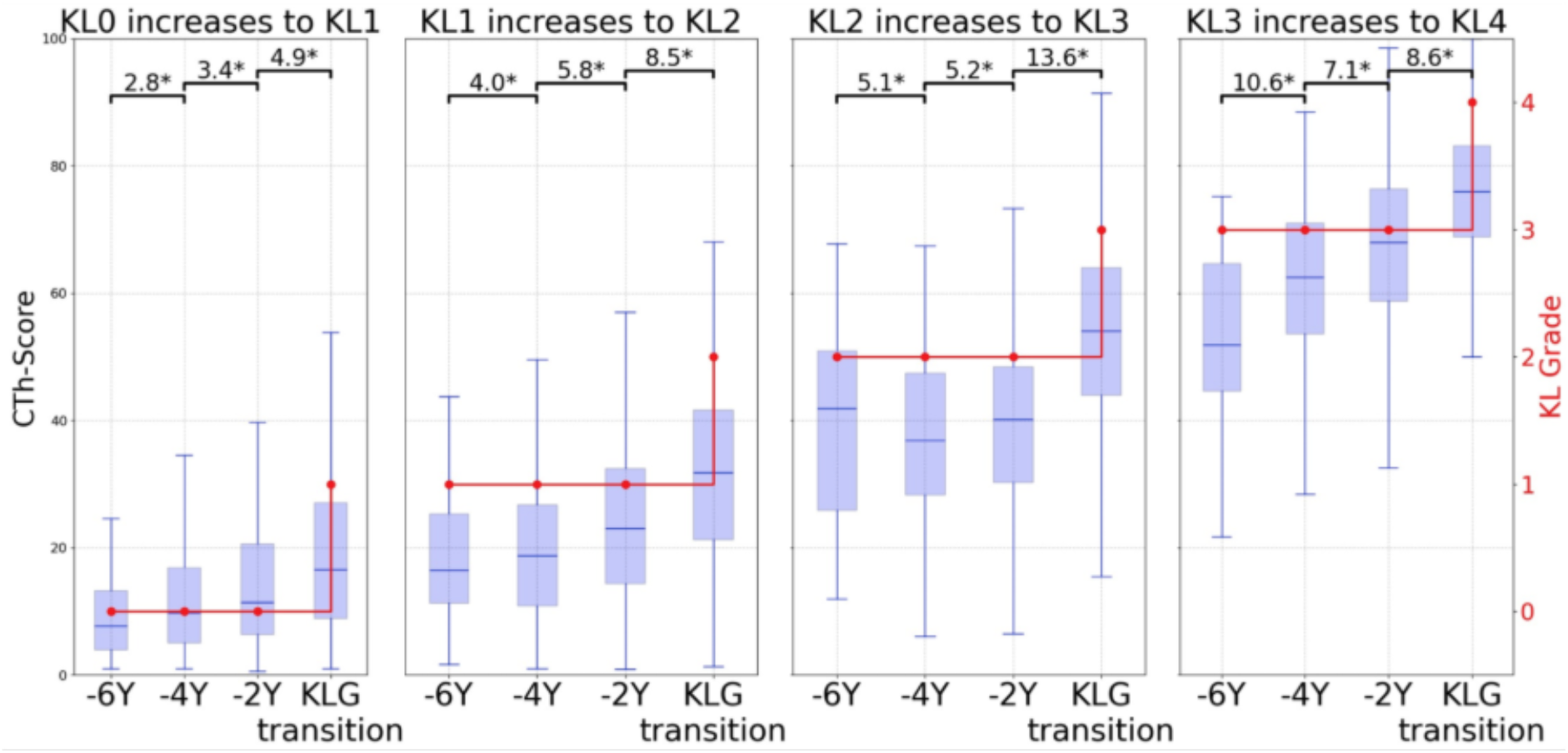
Evolution of CTh-Scores prior to radiographic OA progression. Four different KL grade increases to grade+1 are considered. KL2 is the threshold for radiographic OA. ‘KLG transition’ represents the timepoint where the subject knees have been graded KL+1. The CTh-Scores of these knees are reported 6, 4 and 2 years before these transitions. The important range of CTh-Score in the KL2 and KL3 reflects the diversity of cartilage degeneration severity in the active states of the disease. At all stages of the disease (including pre-radiographic OA) the CTh-Scores consistently increased between every timepoint starting 6 years prior the KLG transition.

### Statistical analysis

Due to the non-normal distribution of disease severity in the OAI and external datasets, Spearman correlations were performed, and a rank-based inverse normal transformation was applied to calculate the 95% confidence interval[28]. In figure 5, due to the non-normal distribution of differences between the groups’ timepoints, a paired Wilcoxon signed-rank test was employed to assess the significance of changes in the CTh-Score between consecutive timepoints leading up to the grade transition[29]. To control for multiple comparisons, the Bonferroni correction method was applied, as it provides a conservative adjustment.

## Results

### Segmentation of bone and cartilage

The segmentation model achieved high performance, with Dice coefficients of 89.6 ± 2.4 for femoral cartilage, 85.8 ± 4.0 for tibial cartilage, and 98.5 ± 0.27 and 98.7 ± 0.31 for femoral and tibial bone, respectively (N = 507). We adopted the same cross-validation testing protocol as a recent study [30] using the same training dataset and our model produced comparable segmentation results [30]

### Repeatability of CTh-Maps and CTh-Score

For the CTh-Maps, the accuracy was –0.002 ± 0.018 mm, and the precision 0.141 ± 0.031 mm. The repeatability across all maps was excellent, with an ICC of 0.98 ± 0.01.

The CTh-Score demonstrated excellent repeatability, with an ICC of 0.99, and a smallest detectable change of 4.7, which corresponds to 4.7% of the CTh-Score’s range.

### Generalizability of CTh-Score

As depicted in Figure 3, the CTh-Score demonstrated sensitivity to OA severity and generalizability across these datasets. The CTh-Score showed a significant increase between successive KL grades in all cohorts. Notably, this sensitivity was evident even at pre-radiographic disease states (KL grade 0 and 1), where radiographic OA is not yet declared. This finding held true for the external dataset as well, despite the use of a different imaging modality (arthro-CT) and patient population, highlighting the CTh-Score’s ability to reflect cartilage status in relation to OA severity across varied conditions.

### Validity of CTh-Score

#### CTh-Score trajectories in the OAI cohort over 8 years

Over the 8-year study period, the CTh-Score showed an increase in 95% of the participant’s knee (4,696 out of 4,953) from the OAI cohort. Among the minority of subjects with a decrease in CTh-Score, the median reduction was –1.6, with an interquartile range of [–3.5, –0.6]. The changes in CTh-Score over 8 years (DCTh-Score) revealed three distinct trends relative to the baseline CTh-Score. First, a relative slow progression that accelerated from a baseline CTh-Score of 0 to 20, with the median DCTh-Score rising from 3.0 to 15.0. Second, a relatively stable high increase from a baseline CTh-Score of 20 to 60 (with the median CTh-Score relatively constant from 15.0 to 15.6). Third, the increase decelerated from a baseline CTh-Score of 60 to 90, with the median DCTh-Score dropping from 15.6 to 1.1.

#### Relationship with cartilage loss and global disease severity

In the OAI MOAKS sub-cohort at baseline, the correlation between the CTh-Score and total size of cartilage loss was strong (r = 0.81 [0.79,0.82]; p<10^-9^; n = 2671). Importantly, it is superior to the correlation between the KL grades and the total size of cartilage loss (r = 0.68 [0.66,0.70]; p < 10^-9^; n = 2671).

#### Sensitivity to disease progression

Considering the OAI imaging timepoints of baseline (0 months), 24 months, 48 months, and 96 months, a total of 289 knees transitioned from KL grade 0 to grade 1, 317 from 1 to 2, 352 from 2 to 3, and 257 from 3 to 4. At all stages of the disease (including early stages), the CTh-Scores increased significantly between every timepoint, starting 6 years prior to the KL grade transitions.

## Discussion

This study introduces a novel approach to analyzing cartilage in osteoarthritis imaging. The proposed framework offers an automatic, highly sensitive, reproducible, quantitative evaluation of OA cartilage severity, extensively validated against established standards for expert assessment of cartilage loss (MOAKS grading) and OA global severity (KL).

By leveraging recent advances in automatic segmentation, we have generated standardized cartilage thickness images (CTh-Maps), revealing diverse cartilage degeneration patterns, which would be obscured by averaging methods using ROIs. The automatic pipeline for CTh-Maps production demonstrates excellent reproducibility, delivering accuracy and precision comparable to the reference standard of manual processing in test-retest analysis. This method is further adaptable to different datasets and imaging modalities, requiring only minor adjustments for bone and cartilage segmentations. The rapid (1 min per scan) and automatic image processing enabled the analysis of the entire OAI cohort, creating a large dataset of bilateral CTh-Maps for 4796 subjects across eight timepoints, totaling more than 47,000 CTh-Maps. These images offer valuable insights into the cartilage degeneration patterns and open the door for a range of further analyses.

Furthermore, the detailed information on CTh patterns contained in the CTh-Maps enabled the development of the CTh-Score, a continuous measure of OA cartilage severity. Interestingly, the ability for a convolutional neural network to quantify cartilage degeneration was developed through supervised training, using the KL, a marker of global OA severity, as the target. It is important to note that KL was not used as a variable to be predicted, but rather as a weak supervisory signal to guide the learning of a continuous, morphology-based severity score. The objective of this study was therefore to derive the CTh-Score as a new quantitative construct of osteoarthritic cartilage severity, rather than to model or reproduce the ordinal KL grades themselves. While the KL provides a relatively coarse grading, it serves as a robust proxy for cartilage degeneration severity: higher KL grade values generally correspond to more pronounced cartilage degeneration. In contrast, traditional cartilage loss metrics, such as joint space width or regional average measures of CTh on MRI, are poor predictors of lesion severity due to their high inter-subject variability, which can lead to significant overlap between healthy and OA cartilage. Additionally, cartilage lesions manifest as complex patterns of both thinning and thickening, rather than mere cartilage loss, which information cannot be effectively conveyed by joint space width or regional average measures of CTh. Although radiographic severity provides a coarse training signal, a key strength of the CNN is the ability to extract more nuanced information from input image patterns. This phenomenon has already been shown in OA classification using knee radiographs to predict KL, where authors found that the probability distribution across KL classes provided a more refined estimation of disease severity[31]. Just as the probability outputs of a classification network convey more detailed information than class labels themselves, the CTh-Score captures more granular detail about the severity of OA cartilage than the KL used for training.

The CTh-Score effectively reflects OA cartilage severity, demonstrating a strong monotonic correlation with both the expert evaluations of cartilage loss (derived from MOAKS gradings) and radiographic disease severity as reflected by the KL. It correlates more strongly with the MOAKS gradings than the KL grades, indicating greater sensitivity to cartilage degeneration than radiographic grading. Furthermore, the CTh-Score exhibits excellent reproducibility and a smallest detectable difference of 4.7%.

Importantly, the CTh-Score demonstrated generalizability, as it was successfully applied to an external CT-arthrography dataset without any retraining. Despite the fundamental differences in how cartilage is visualized and delineated in arthro-CT compared to MRI, owing to the distinct contrast mechanisms and spatial resolutions, the CTh-Score maintained consistent associations with osteoarthritic severity as reflected by the KL grade. This finding underscores the robustness of the framework and its ability to capture patterns in the CTh-Maps that generalize across imaging modalities and populations. However, while these results are encouraging, the influence of imaging modality on CTh-Score estimation warrants further investigation to quantify potential systematic differences between MRI- and CT-derived cartilage thickness measurements.

An analysis of CTh-Score trajectories in the OAI provides interesting insights in the ability of MRI to quantitatively detect cartilage degeneration. Critically, the CTh-Score consistently detected cartilage degeneration progression between timepoints, up to 6 years before radiographic progression became apparent, regardless of the OA stage. This underscores the sensitivity of the CTh-Score to the progression of cartilage degeneration and thereby its potential as a tool for early detection and monitoring of OA.

Furthermore, CTh-Score trajectories reveal substantial inter-subject diversity. Some knees exhibit rapid progression across nearly the entire CTh-Score range during the OAI study, while others show minimal progression despite advanced lesions. This variability aligns with a previous study on cartilage thickness evolution, which also reported diverse trajectories over 8 years in the OAI[32]. However, a general three-phase trend emerges: an initial period of relatively slow progression, followed by an accelerated phase corresponding to active cartilage degradation, and finally a deceleration in later stages when lesions are already severe and have limited capacity for further progression. This dynamic, consistent with cartilage pathophysiology[33], underscores the importance of detecting early cartilage degeneration, when the rate of progression is still low. This early stage represents a window of opportunity[34] for interventions aimed at slowing or reversing cartilage degradation. The CTh-Score, with its proven sensitivity to the progression of OA cartilage severity, serves as a valuable tool for identifying this window during the screening phase of clinical trials.

Although the underlying architectures (U-Net and ResNet) are standard, the originality of this work lies in the development of a fully automatic, end-to-end framework that transforms raw MRI images into standardized CTh-Maps and a quantitative, continuous score of osteoarthritic cartilage severity.

This study has certain limitations that should be acknowledged. When applied to a new dataset, the segmentation model requires retraining using ground truth segmentations. This registration method performed well across knees from the OAI dataset, including KLG4 knees with severe bony deformities, it may not be fully generalizable to severe post-traumatic deformities or rare congenital and developmental abnormalities (e.g., achondroplasia).

While the CTh-Score provides a simple yet effective one-dimensional metric of osteoarthritic cartilage severity, it cannot fully capture the complete information contained in CTh-Maps. Future work should explore the extraction of degeneration patterns from CTh-Maps, particularly those associated with pain, biomechanics, or specific progression phenotypes. Although the CTh-Score demonstrated strong performance despite being derived from the ordinal and relatively coarse KL grade, a more granular representation of cartilage degeneration may be achievable using refined ground truths. Histology or arthroscopic assessments, for instance, could provide more specific biochemical or structural cartilage information. Future work will also include direct comparisons between the CTh-Score and quantitative MRI biomarkers such as T2 mapping. In addition, while the framework has demonstrated scalability and generalizability across imaging modalities, further investigation is warranted to quantify the impact of imaging protocol variations, scanner differences, and sequence dependencies.

In conclusion, this study establishes a robust and automatic framework for generating CTh-Maps, enabling detailed analysis of cartilage thickness patterns in OA. The CTh-Score derived from these maps provides a sensitive and reproducible tool to quantitatively assess OA cartilage severity, offering insights into disease progression that surpass traditional assessments. These findings highlight the potential of CTh-Maps and of the CTh-Score to gain insights into the different patterns of CTh progression leading to an earlier OA detection, better disease monitoring, and the identification of therapeutic windows.

## Data Availability

All data produced in the present study are available upon reasonable request to the authors

## Acknowledgements

This work was funded by the Swiss National Science Foundation, Switzerland (SNSF Grant #CRSII5_177155 & CRSII--222725). The authors thank the Osteoarthritis Initiative (OAI) investigators, clinical staff and participants at each of the clinical centres and at the coordinating centre for their important contributions in acquiring the publicly available clinical and imaging data. The OAI is a public–private partnership comprising five contracts (N01-AR-2-2258; N01-AR-2-2259; N01-AR-2-2260; N01-AR-2-2261; N01-AR-2-2262) funded by the NIH and conducted by the OAI Study Investigators. Private funding partners of the OAI include Merck Research Laboratories, Novartis Pharmaceuticals Corporation, GlaxoSmithKline, and Pfizer. Private sector funding for the OAI is managed by the Foundation for the NIH. JF and PO equally contributed to this work and should be considered as co-last authors.

